# A pilot study of the quality of care of atrial fibrillation in Irish general practice

**DOI:** 10.1101/2023.06.12.23291272

**Authors:** Sarah McErlean, John Broughan, Geoff McCombe, Ronan Fawsitt, Mark Ledwidge, Walter Cullen, Joe Gallagher

## Abstract

**Aim:** The aim of this pilot study is to develop and test a quality of care score for patients with permanent atrial fibrillation (AF) in general practice, and identify the patterns of quality of care using this score.

We developed a quality of care score based on the Atrial Fibrillation Better Care pathway recommended by the European Society of Cardiology and the European Heart Rhythm Association guidelines. This is a 14-point score that we have termed the MAGIC score(Management of Atrial Fibrillation in Integrated Care and General Practice).

**Methods & Results:** An observational cross-sectional pilot study was undertaken. Proportionate sampling was used across 11 practices from the Ireland East practice-based research network. The GPs completed a report form on each patient provided by the research team by undertaking a retrospective chart review.

11 practices participated with a total number of 1855 patients with AF. We received data on 153 patients.

The main findings were that:

No patient met all 14 guideline based recommendations Mean MAGIC score was 11.3

Points were most commonly deducted because the creatinine-clearance and HAS-BLED score were not recorded and the patient was not on the correct dose of oral anti-coagulation.

**Conclusion:** This study demonstrates the feasibility of using a quality of care score to measure quality of AF management in general practice. This scoring system; which is based on internationally recognised quality of care markers, highlights key areas that can be targeted with intervention to improve care.

## Introduction

Atrial fibrillation is the most common sustained cardiac arrhythmia in adults across the world and poses a significant burden to patients, physicians and healthcare systems globally.^1^ The prevalence of AF is rising due to an ageing population, more vigorous searching for undiagnosed AF and the increasing burden of co morbidities.

Despite progress in the management of AF, it is still a leading cause of stroke, heart failure and cardiovascular morbidity and mortality. ^2^ AF is also associated with reduced quality of life (QoL), depression, cognitive decline and hospitalisation. ^2,3^ Adherence to guideline directed therapy can improve outcomes in AF and yet there are wide variations in compliance to these guidelines.^4,5^

The latest ESC guidelines highlight that patients with AF have a broad range of adverse cardiovascular outcomes and the approach to management should be comprehensive and holistic. This can be achieved using the Atrial Fibrillation Better Care (ABC) pathway. ^1^ This ABC pathway streamlines integrated care of AF patients across all levels and specialities of healthcare and has shown significant benefits and improved clinical outcomes.^1,6–8^

Given the prevalence of AF, optimal management is paramount to improve quality of life and reduce overall impact of AF on the health and social care system. ^9^ Measuring quality and identifying opportunities for improvement is an essential part of optimal AF management. ^1^ Guidelines are rarely absorbed organically into clinical practice sufficiently enough to create a meaningful change. Additional QI interventions are often needed and usually a combination education, decision support, ongoing audit and feedback and organisational change are necessary for a sustained effect. ^10^Good quality data acquisition, audit and feedback is essential to highlight areas in need of intervention and monitor progress. We know that audit and feedback can lead to important improvements in clinical care and drive behaviour change. ^11,12^

The aim of this pilot study is to develop and test a quality of care score called the MAGIC score (Management of Atrial Fibrillation in Integrated Care and General Practice) for patients with permanent atrial fibrillation in general practice, and identify the quality of care in general practice using this score.

## Methods

### Development of quality of care score

The quality of care score is based on the ABC pathway and the European Heart Rhythm Association guidelines and quality indicators. ^2,13^ The score was developed by an expert panel of general practitioners using EHRA criteria that were 1. Relevant to general practice 2. Extractable from the patient record 3. Amenable to a quality improvement intervention.

The final 14-point score is outlined in Table 1.

**Table 1:** MAGIC* Score for quality of care in general practice.

|  |  |
| --- | --- |
| 1 | Appropriately prescribed anticoagulation according to CHA2DS2-VASc score |
| 2 | Patients with permanent AF who are not inappropriately prescribed antiarrhythmic drugs |
| 3 | TSH evaluation since diagnosis |
| 4 | Fasting glucose / HbA1C measurement since diagnosis |
| 5 | Renal function measured since diagnosis |
| 6 | Echocardiography performed since diagnosis |
| 7 | BP measurement since diagnosis |
| 8 | CHA2DS2-VASc score recorded since diagnosis |
| 9 | HAS-BLED score recorded since diagnosis |
| 10 | Creatinine Clearance calculated since diagnosis |
| 11 | Prescribed correct NOAC dose |
| 12 | On anti-platelet only if indicated |
| 13 | Weight recorded |
| 14 | BP < 140 / 90 mmHg |

**\*MAGIC = Management of Atrial fibrillation in General practice and Integrated Care**

The first 7 points were similar to those which were included in a recent randomised control trial comparing nurse-led care to usual care which assessed adherence to 7 guideline based recommendations. ^14^

### Pilot study

An observational cross-sectional pilot study was undertaken to determine the quality of care in general practice of atrial fibrillation using this score.

### Setting

The study involved general practices (n=14) participating in a research network on the east coast of Ireland. GPs who agreed to participate (n=11) returned a signed consent form and the total number of patients with permanent AF in their practice if they were happy to participate. Proportionate sampling to generate a sample size for individual practices.

A random number generator was used to identify patients from their individual list in each practice to include in the study. A paper case report form was used to collect defined anonymised data on these patients. The report forms were returned to the research team in UCD. This data was then cleaned, pooled and inputted onto a secure database.

### Participants

All practices in the Ireland East practice-based research network were invited to participate.

They identified patients in the practice with permanent AF.

Inclusion Criteria:

- Age ≥ 18 years
- Permanent AF detected on ECG, Holter recording or event recorder
- Active in practice software and has attended the practice in the last 24 months

Exclusion Criteria:

- No electrocardiographic objectified AF

### Study Size

As this was a pilot study a formal sample size was not calculated. ^15^ Based on previous work it was estimated that 12 per group would be sufficient. ^16,17^We estimated that 80% of the practice would complete the study (11 practices). Therefore we sought to recruit a minimum of 132 patients using proportional sampling.

### MAGIC Score

Each participant file was reviewed using the MAGIC score and points awarded if the relevant component was present. Points were not awarded if the information was not recorded or could not be determined.

### Statistical Methods

RedCap ^18^ was used for data storage and analysis. The SPSS programme was used for statistical analysis.

## Results

11 practices consented to participate in this pilot study with a total number of 1855 patients with AF. We received data on 153 patients proportionally sampled across these 11 practices.

The demographic details can be seen in Table 2.

**Table 2.**
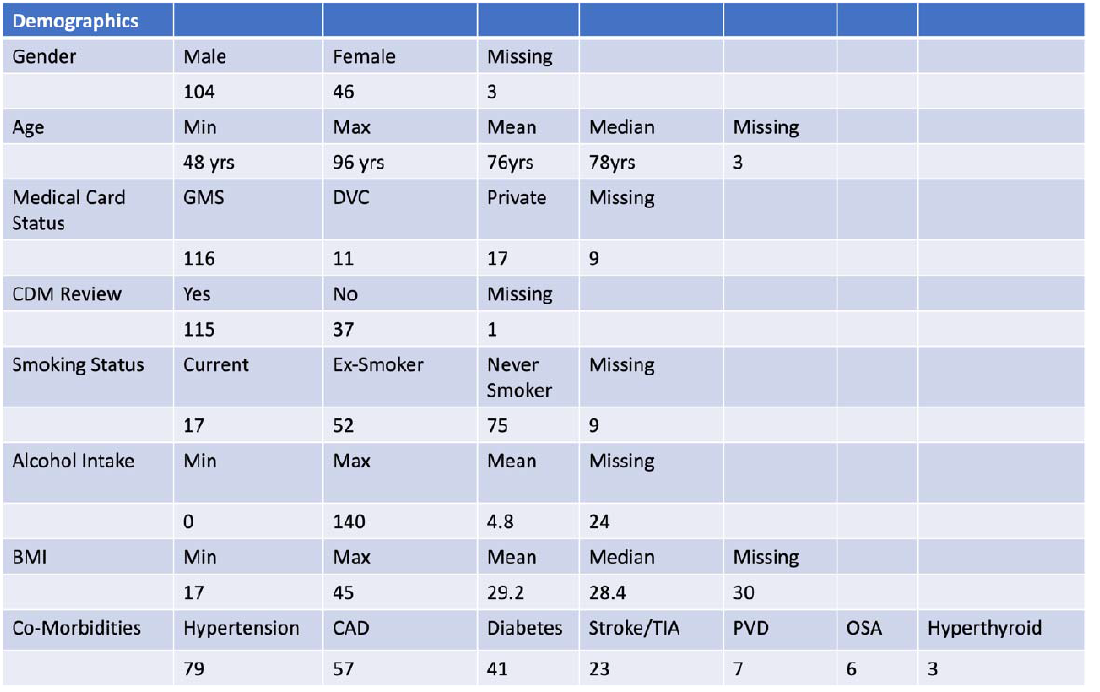

Most patients – 68% were male (N= 104) and the mean age was 76 years. The majority of patients (75%) had a “General Medical Service” (GMS) card which entitles patients to free GP care in Ireland. These are awarded to all patients over 70 years, under 6 years of age and those below a certain income level based on means testing. These patients are also eligible for a biannual structured chronic disease management (CDM) review which was introduced in March 2020. This CDM review is not available to patients without a medical card. Most patients had a CHA2DS2-VASc score calculated (76%) and most patients (86%) were on oral anticoagulation (OAC). The most commonly used OAC was Apixaban (52.3%). Of the 21 patients not on OAC, this was only appropriate in 4 of them based on their CHA2DS2-VASc score. See Figures 1 & 1a.

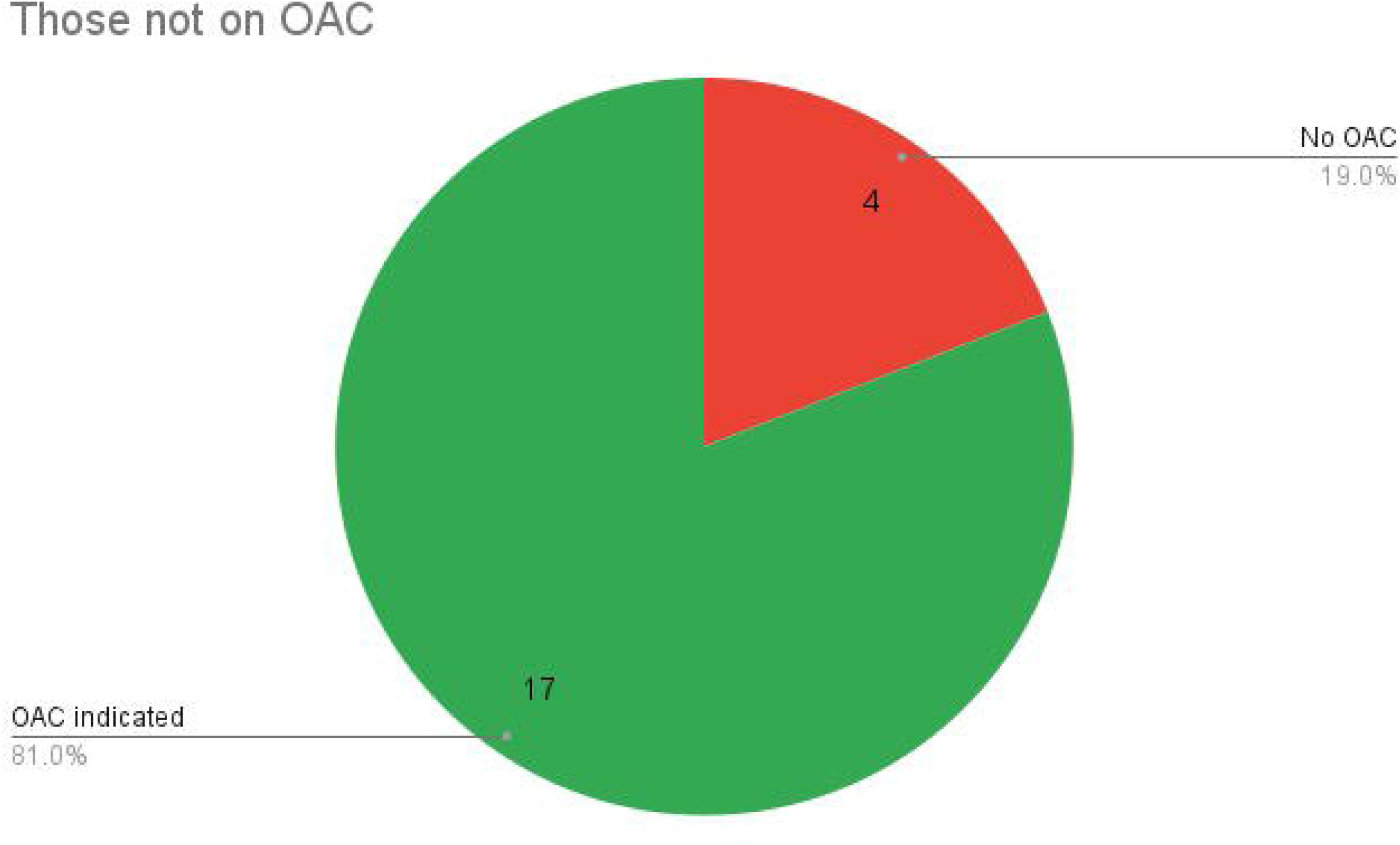

The mean heart rate was 74bpm (rang 42-124bpm) and the mean blood pressure was 132 / 77 mmHg (systolic BP range 90-207mmHg, diastolic BP range 51-103mmHg). 75% of patients were on rate controlling medication. 99.3% of patient had a HR < 110 bpm. Bisoprolol was the most common medication used (54%). 12% of patients were prescribed anti-arrhythmic drugs despite being in permanent atrial fibrillation. See Figure 2 & 3.

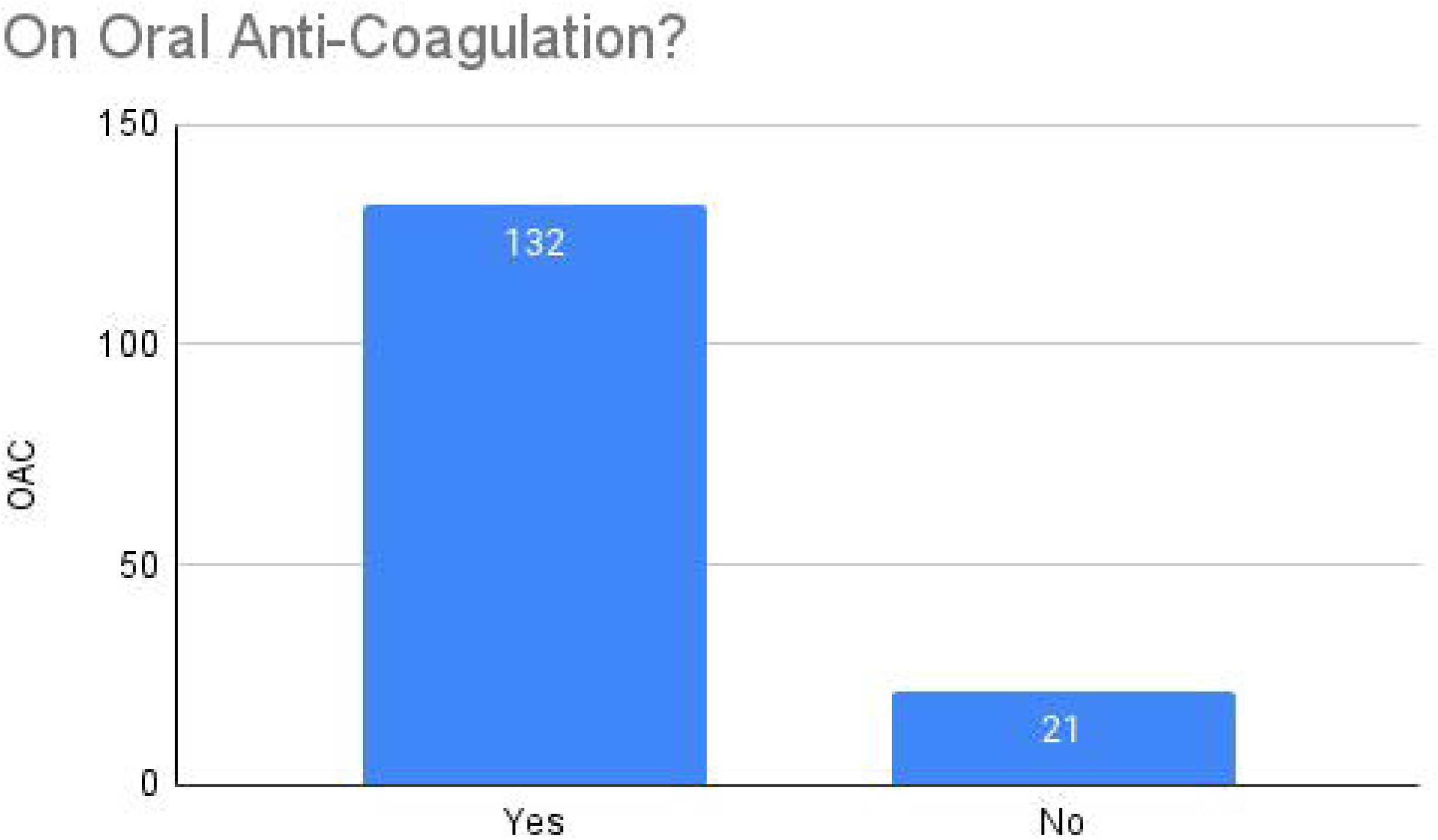

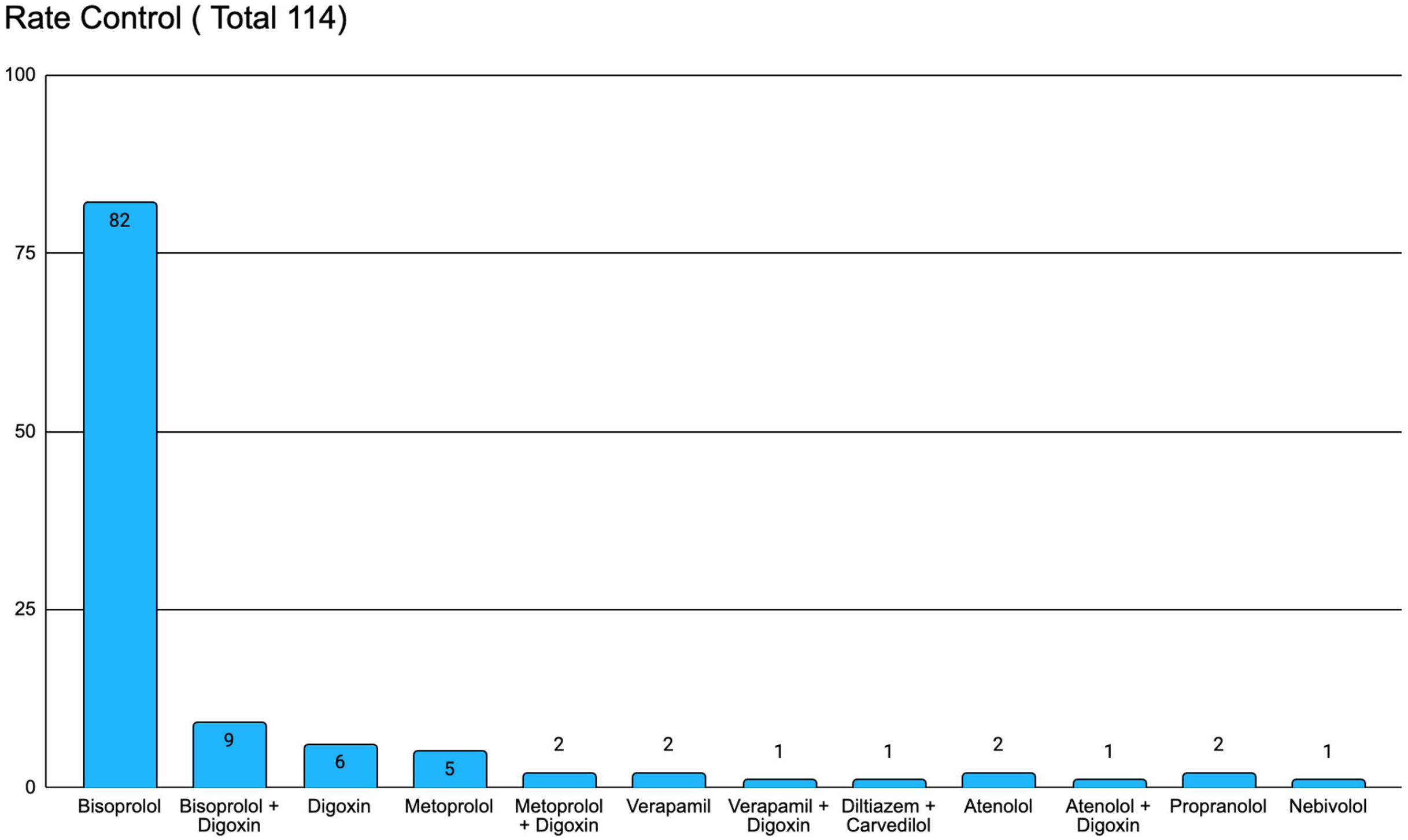

The mean BMI was 29.2 kg/m2 (Range 17-45 kg/m2) with 36.4% being above 30kg/m2. 45% were current or ex-smokers, 49% never smoked and the mean weekly alcohol intake was 4.8 units. Hypertension was the most common co-morbidity recorded followed by coronary artery disease.

Using the MAGIC score no patient met all 14 guideline based recommendation. Only 13.8% met 13 based recommendations. As illustrated on the graph there is a sharp decline after a score of 10. The mean score was 11.3 points. (Figure 4) Points were most commonly deducted because the creatinine-clearance and HAS-BLED score were not recorded and the patient was not on the correct dose of OAC. (Figure 5)

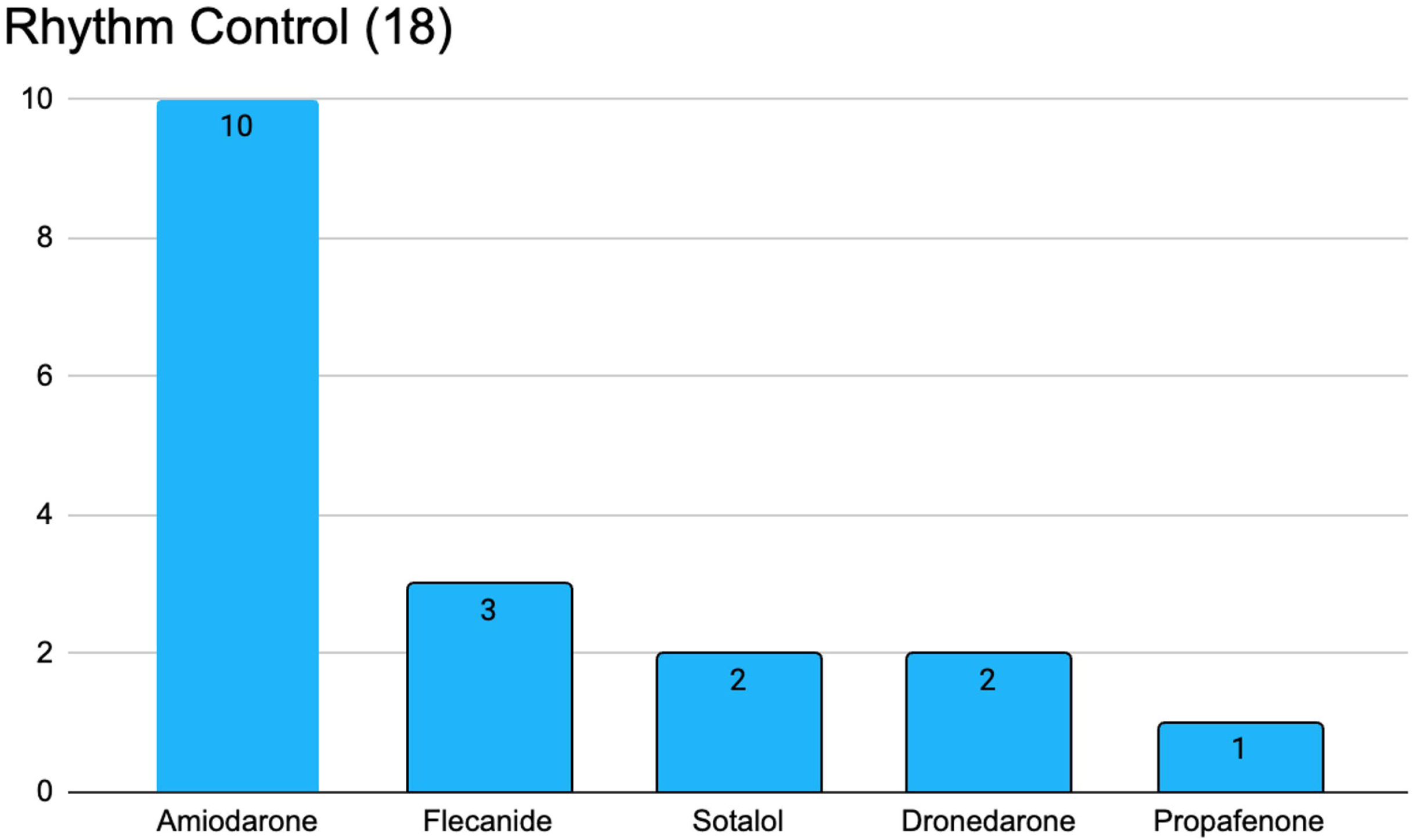

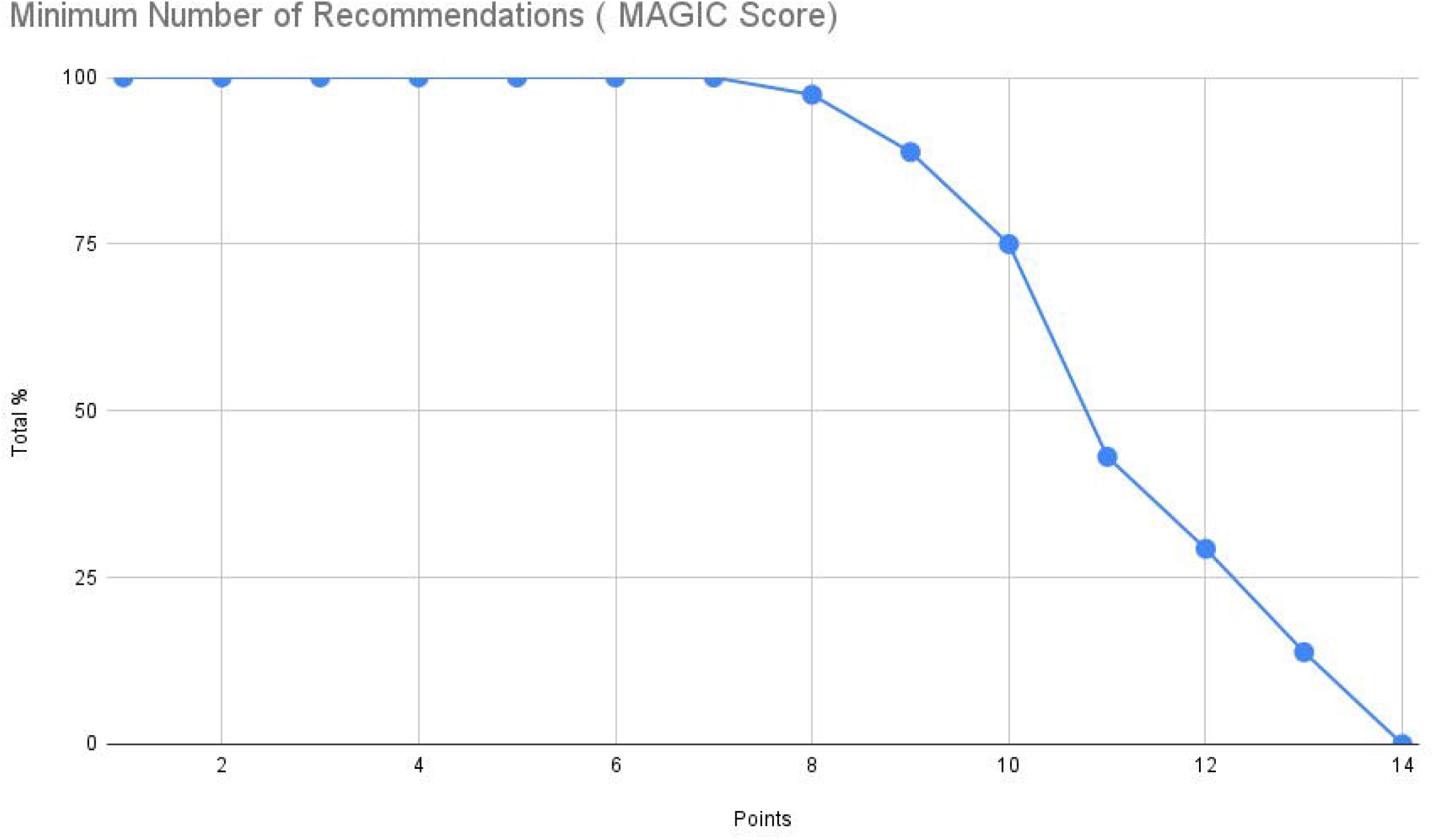

Only 29 patients had their creatinine clearance calculated despite the fact that nearly all patient had their renal function checked (150 patients) since starting on OAC. 52 patients were either taking Rivaroxaban, Edoxaban or Dabigatran which necessitates this calculation to ensure correct dosing.

## Discussion

### Key Results

This study demonstrates the patterns of quality of care of atrial fibrillation in general practice and the feasibility of using a quality of care score. It shows that the majority of the makers of quality of care described were adhered to. A number of areas that are amenable to quality improvement interventions were identified. By measuring quality indicators at regular intervals we can continue to assess and improve patient care.

A recent RCT that found that compliance to 7 guideline based recommendation was 61% under specialist nurse led care and 26% in usual care. We found that 59.5% of patients met these same 7 guideline based recommendation in the general practice cohort that we assessed. (Figure 6) Most patients in our study had a CDM review involving attendance with a general practice nurse and GP review at 6 monthly intervals. Previous studies have shown that structured care in general practice are important for improving outcomes.^6,19,20^ International guidelines for clinical AF management have recommend integrated care as the leading approach to manage AF resulting in improved clinical outcomes. ^1,6–8,20^Part of integrated care is the adherence to guideline directed therapy.^9^

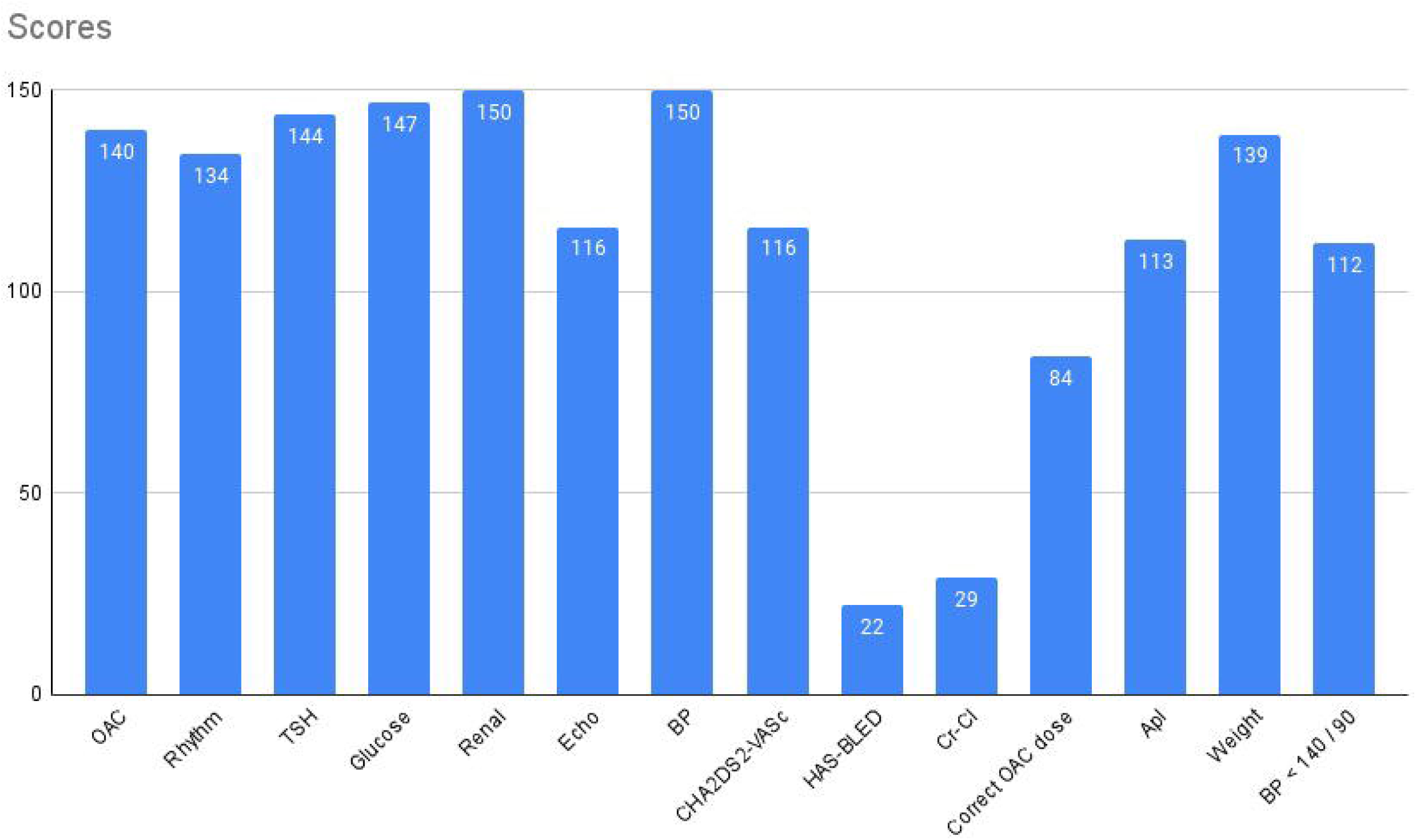

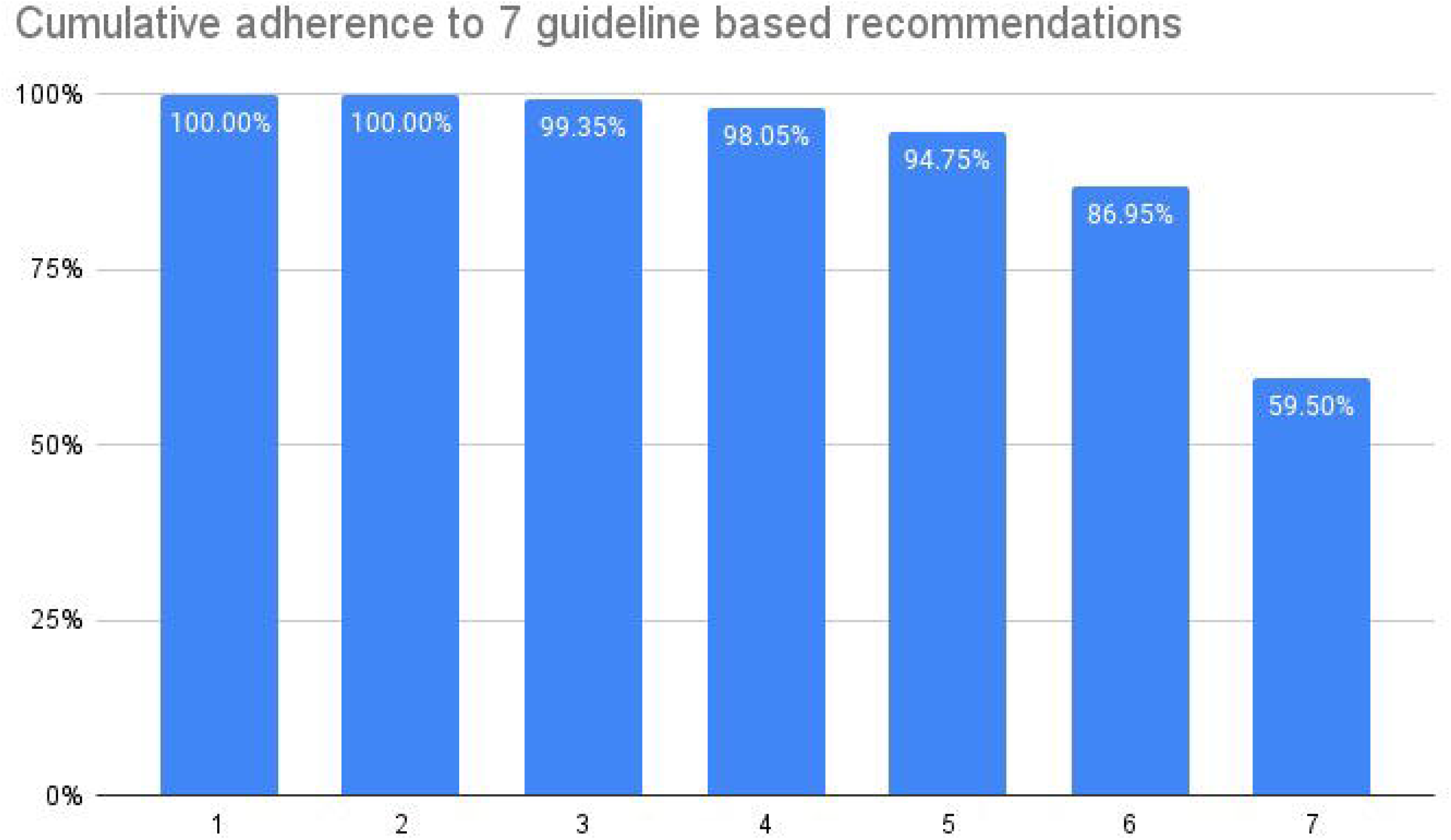

Most patients (86%) in this pilot study were on OAC. Based on the patient’s CHA2DS2-VASc score of 1 in a male or 2 in a female, OAC was indicated in 17 of the 21 patients not prescribed OAC. Of those taking NOACs, 80% were on the correct NOAC dose. The areas of the MAGIC score that had the poorest results were the recording of a HAS-BLED score and recording of creatinine clearance which are essential components of OAC management. These are amenable to electronic recording and calculation. 22 patients in this study had a HAS-BLED score recorded. Carrying out an individual stroke and bleeding risk assessment in a key component of AF management. Using the HAS-BLED score has been shown to improve appropriate OAC prescribing. ^21^ In the mAFA-II trial, using the HAS-BLED score to dynamically assess bleeding risk was associated with increased prescribing of OAC, reduced bleeding events and mitigated modifiable risk factors for bleeding. ^22^ Poor compliance with assessment of bleeding risk means that those at higher risk of bleeding are not identified and therefore not managed correctly.

In terms of better symptom control, the focus is on rate control in those with permanent atrial fibrillation in general practice. Patients in this study had a mean heart rate of 74 bpm and only 1 patient with a heart rate of > 110bpm was identified. Beta blockers with the addition of Digoxin were the most common rate controlling medications used. AADs were potentially inappropriately prescribed in 18 patients. There is no role for long term AAD in those with permanent AF (i.e. where no attempt to restore sinus rhythm is planned) ^2^ and these patients may have been exposed to the side effect profile of these drugs unnecessarily unless being used for other indications also.

Managing comorbidities is an important component of atrial fibrillation care. AF often results from a combination of underlying interacting factors leading to atrial remodelling/cardiomyopathy including cardiovascular disease, cardiovascular risk-factor burden, co morbidities and an unhealthy lifestyle. ^1^ Hypertension is the most common aetiological factor for AF. The mean BP in in this cohort was 132/77mmHG with 24.8% patients having a BP >140/90mmHg. The mean BMI was 29.2 kg/m^2^ which is classed as overweight and nearly obese. Obesity not only increases the risk of AF but also the risk of ischaemic stroke and death.^1^ Weight loss has been shown to be associated with better atrial fibrillation control. ^23^

A number of interventions have been tested which have been shown to improve quality of care in chronic diseases. The American Heart Association’s (AHA) “Get With The Guidelines” (GWTG) programme has shown to improve quality of care across a number of different cardiovascular diseases and has now been expanded to AF. ^10^ Participating hospitals record patient details on a quality improvement registry and guideline adherence is facilitated using rapid – cycle quality improvement strategies and site specific reporting. ^10^ Sites are provided with performance data every 3 months which enables targeted interventions to be utilised mainly through education and support. ^10^The approaches used includes educational intervention, availability of treatment algorithms and decision support tools.^24^Using the GWTG approach, appropriate OAC prescription in patients with AF improved significantly demonstrating that a high level of guideline adherence is possible. ^24^The IMPACT-AF trial had similar findings. By using a multifaceted and multilevel educational intervention, the appropriate prescription for OAC significantly increased. ^25^

These QI interventions have also been used successfully in primary care. It has been shown that a combination of education and specialist support resulted in an improvement in OAC prescribing and the number of patients undergoing both stroke and bleeding risk assessments. ^19^

A group of general practitioners in the UK have developed an interactive dashboard to provide feedback on quality in AF care. They found that the dashboards were an acceptable and feasible way to communicate a summary of AF care. ^26^ The prescribing section was rated as one of the most useful by the pilot group of GPs. Translating these approaches to the range of quality indicators in the MAGIC score could prove useful in improving management of atrial fibrillation in general practice.

### Strengths and limitations

The MAGIC score enables identification of gaps in AF care and guideline adherence. This score can be used for ongoing audit and feedback which is a key part of quality improvement. This score is easy to use and could be replicated across other general practice sites as the information we collected is readily available within the electronic health record (EHR). This type of score readily identifies areas that can be targeted with an intervention to improve quality of care. It is based on internationally recognised quality of care markers. It involved proportionate sampling across 11 practices providing a representative sample. A number of limitations apply to this study also. We assessed if patients had ever had the relevant investigations since diagnosis of AF but did not record the time since diagnosis to performance of the relevant investigation. We also did not record the time in target range (TTR) for patients on warfarin. For those not on anticoagulation or incorrect doses according to guidelines we did not ascertain if there were reasons for the non-use or use of different NOAC doses.

This MAGIC score has not been validated and we do not know if a higher score translates to better clinical outcomes, although other work in this area has demonstrated that adherence to guidelines is beneficial for patients.^4,5,20^ However overall this was a pilot study to develop and pilot the score and this was outside the scope of the project.

## Conclusion

The ESC highlight the importance of measuring quality with quality/performance indicators to identify opportunities for improvement.^1^ We propose that the MAGIC score can be used for this purpose. In this pilot study, the quality of AF care in Irish general practice is good and comparable with other work in this area. The MAGIC score has highlighted some key areas that can be targeted with intervention to improve quality of care. Decision support tools, educational interventions, and visual summaries such as dashboards have all been shown to be effective QI strategies to improve guidelines adherence and will help inform future work.

## Data Availability

All data produced in the present study are available upon reasonable request to the authors

